# Comparative Evaluation of SARS-CoV-2 IgG Assays in India

**DOI:** 10.1101/2020.08.12.20173856

**Authors:** DBT India Consortium for Covid-19 Research, Bhatnagar Shinjini

## Abstract

IgG immunoassays have been developed and used widely for clinical samples and serosurveys for SARS-CoV-2. We compared the performance of three immunoassays, an in-house RBD assay, and two commercial assays, the Diasorin LIAISON SARS-CoV-2 IgG CLIA which detects antibodies against S1/S2 domains of the Spike protein and the Zydus Kavach assay based on inactivated virus using a well-characterized sera-panel. 379 sera/plasma samples from RT-PCR positive individuals >20 days of illness in symptomatic or RT-PCR positivity in asymptomatic individuals and 184 pre-pandemic samples were used. The sensitivity of the assays were 84.7, 82.6 and 75.7 respectively for RBD, LIAISON and Kavach. Kavach and the in-house RBD ELISA showed a specificity of 99.5% and 100%, respectively. The RBD and LIAISON (S1/S2) assays showed high agreement (94.7%;95%CI:92.0,96.6) and were able to correctly identify more positives than Kavach. All three assays are suitable for serosurveillance studies, but in low prevalence sites, estimation of exposure may require adjustment based on our findings.

## Introduction

Nucleic acid-based diagnostic tests like RT-PCR have shown considerable sensitivity and specificity for detection of active SARS-CoV-2 infections and are being used as the primary diagnostic tool. Serological tests, on the other hand, are important tools for estimation of seroprevalence of the disease at a community level. At an individual level, serological evidence may be a correlate of exposure or vaccine response.

Performance characteristics of serological tests determine their utility and interpretation of results. For SARS-CoV2, many tests have been developed but there are limited direct comparisons. Approximately 390 tests for IgG, IgM, IgA and total antibody have been developed across a range of platforms including chemiluminescence, magnetic bead-based assays, microwell ELISA, lateral flow, etc. using different portions of the spike and nucleocapsid proteins as well as whole inactivated virus [1]. Although the nucleocapsid is more abundant and immunogenic, most assays in use or in development have utilized different regions of the spike protein or whole virus as the capture reagent in immunoassays. This is mainly because antispike antibodies are believed to be less cross-reactive based on viral spike homology and are expected to correlate better with neutralizing capacity of convalescent sera. Most of these assays have been developed rapidly, many under emergency use authorization, and hence were evaluated by the developers in a limited set of samples. The performance of these assays in larger sample sets in various real-world setting is necessary to interpret the results of the seroepidemiological studies conducted using these assays.

In this study, we evaluate three serological tests, one in-house ELISA, a commercial ELISA, and a commercial chemiluminescence immunoassay and report their sensitivity based on 379 RT-PCR positive convalescent sera and plasma samples collected from a prospective cohort of COVID-19 positive participants and specificity based on 184 pre-pandemic participants. We also perform head-to-head analyses of their ability to correctly identify IgG positive samples.

## Materials and Methods

### Participants for serological assay comparison

The participants for this study were derived from a longitudinal cohort of COVID-19 positive participants known as the Department of Biotechnology India COVID-19 Consortium cohort, with ongoing recruitment from March 2020 at eight clinical sites in the Delhi-National Capital Region, India. The participants in this cohort are derived from two types of enrollment: i) Suspected COVID-19 patients enrolled at the time of RT-PCR testing at the screening center and ii) RT-PCR confirmed COVID-19 positive patients admitted at one of the clinical sites. The testing by RT-PCR was done at an approved laboratory as per the National Testing Strategy of India [2]. The RT-PCR confirmed COVID-19 patients were followed up at 10-28 days and 6-8 weeks of onset of illness. During the enrollment and follow-up detailed clinical information on the exposure history, clinical features and comorbidities were documented. Venous blood samples are collected, transported, processed and stored per protocol [3]. All enrollments were made after an informed consent process and the study protocol was approved by the Institute Ethics Committees of the participating research institutes and hospitals.

The COVID-19 positive reference standard sera panel (n=379) was formed using the sera/plasma samples collected ≥20 days of illness or following RT-PCR positivity. This criterion is in alignment with the UK Medicines and Healthcare products Regulatory Agency (MHRA) to improve comparability [4,5]. The duration of illness for asymptomatic participants was calculated from their date of diagnosis. For symptomatic participants, it was calculated from the date of testing or date of onset of symptoms whichever was the earlier. The COVID-19 negative standard panel was built from sera samples collected in the pre-pandemic period (184 from pregnant women enrolled in a pregnancy cohort) to ensure a clean set of negative samples[4-6].

### Index test methods and alternate reference tests

#### Index test: THSTI In-house RBD IgG ELISA

SARS-CoV-2 RBD protein was diluted to 2μg/ml in PBS pH 7.4 and 50μ/well was coated on maxisorp polystyrene plate (Thermo Scientific). Plates were incubated at 4 °C for overnight. Next day, the wells were washed 3 times with PBST (PBS with 0.1% Tween 20) using 96-well plate-washer (Tecan AG). Two hundred microliter of blocking solution (PBST with 3% skim milk) was added to all the wells. The Plates were incubated at 20°C for 2 hours. After 2 hours of incubation, plates were removed from the incubator and the blocking solution was thrown off. Hundred microliter of diluted test serum or control samples (1:50 dilution in PBST with 1% skim milk) were added to the appropriate wells and the plates were incubated at 20°C. After 2 hours of incubation, plates were removed from the incubator and washed 3 times with PBST. Horse Radish Peroxidase labeled goat anti-Human IgG Fcγ-sp. Tracer antibody (Jackson Immuno Research, Pennsylvania, USA) was diluted 1:5000 in PBST with 1% skim milk and 50 μl of diluted tracer was added in each well of the plates. Plates were incubated in a 20°C incubator for 1 hour. After the completion of the incubation period, plates were washed 3 times with PBST and in each well 100μl of TMB substrate (BD Biosciences) was added and plates were incubated at room temperature for 10 min in dark. Fifty microliter of stop solution (1M H_2_SO_4_) was added in each well and the absorbance was measured using a microplate reader (Biorad, California, USA) at 450 nm with 650 nm as reference wavelength. In each assay, 8 known negative samples with variable background were used as control to calculate the cut-off value. The cut-off value was calculated using the formula: Cut-off = Average OD value of negative control samples + 3* SD of OD value of negative control samples

#### Covid Kavach IgG ELISA

Covid Kavach IgG ELISA was developed by the Indian Council of Medical Research’s National Institute of Virology, and manufactured by Zydus [7]. The test was performed as per manufacturer’s instructions. The kit suggests interpretation of the results by a two-pronged method, based on OD value and P/N (Positive/Negative Ratio). When both read-outs are in agreement, then the sample is considered positive or negative. The manufacturer’s instruction does not mention interpretation for samples with a read-out not in agreement for the two criteria. We considered such results negative.

#### DiaSorin CLIA

The LIAISON SARS-CoV-2 S1/S2 IgG chemiluminescence assay manufactured by DiaSorin was also used for comparison. This test uses S1/S2 antigens to capture specific IgG antibodies. The test was performed as per manufacturer’s instructions, with calibration and positive and negative controls run before each batch of antibody testing as per manufacturer’s protocol [8]. The tests were considered positive when the IgG concentration was ≥15 AU/mL, negative when the concentration was <12 AU/mL and equivocal when the concentration was >12 and <15 AU/mL. Equivocal samples were considered negative for sensitivity analysis.

### Statistical analysis

In the absence of a gold standard for SARS-CoV-2 IgG immunoassays, an alternate reference standard was used for this study, which is SARS-COV-2 RT-PCR positivity with >20 days duration of illness (symptomatic) or >20 days beyond RT-PCR positivity (asymptomatic). While comparing the three assays, we report sensitivity and specificity of each test with the positive and negative reference standards as defined. In addition, we performed a head-to-head analysis for agreement between these methods. We estimated the global agreement calculated as the sum of the number of positives by both methods and number negative by both divided by the total number of samples. We estimated the bias index defined as the difference in the proportion of positivity for any bias between the methods to check whether one method was superior to the other in correctly identifying positive samples. As the prevalence of positives and negatives may play a role in the interpretation of the kappa statistic, we report the prevalence index defined as difference between probability of positives and probability of negatives. We finally report the kappa statistic adjusted for prevalence and bias known as prevalence and bias-adjusted kappa [9]. We then compared the sensitivity of the three candidate tests across different periods of illness. All analyses were done using the STATA-SE-15 software (Texas, USA) and the Kappa coefficient and related indices were estimated using Cohenkap package for STATA [10].

## Results

### Samples defined by an Alternate Reference Standard used for comparative evaluation

The reference true positive sample panel consisted of 379 samples from 368 participants; 11 of whom provided two samples at different time-points. The distribution of the duration of illness was bimodal owing to the design of the cohort from which the samples were derived. The means of the sampling window distributions are 23.5 and 49.3 days respectively (Supplementary figure 1). Most samples (83.7%) were obtained from symptomatic individuals. Nearly half of the samples were sera, rest were plasma. The reference negative panel consisted of 184 pre-pandemic samples collected before September 2019.

### Comparison of the sensitivity and specificity of candidate assays

Among the three candidate tests, RBD IgG ELISA demonstrated the highest sensitivity (84.7; 95%CI: 80.6 - 88.1) and Zydus Kavach the least (75.7; 95%CI: 71.0 - 79.9). Zydus Kavach is interpreted as positive when both test parameters were positive based on cut-Off and P/N ratio. Six samples were indeterminate in Zydus Kavach test; and 25 samples were positive only by one condition (Cut-off, P/N ratio) by Zydus Kavach. Seven samples were reported as indeterminate by DiaSorin CLIA. When Zydus Kavach was interpreted as positive with one of the two criteria, 25 additional samples were identified as positive, improving the sensitivity to 81.8%. The sensitivities of the tests did not change with the increase in the duration of illness beyond 20 days (Supplementary table 5). The specificities of Zydus Kavach and RBD ELISA were 99.5 and 100% respectively (Table-1, Figure-1). The specificity of DiaSorin could not be evaluated due to limited availability of pre-pandemic negative sera.

**Table 1:** Sensitivity (%) and specificity (%) of Zydus-Kavach, DiaSorin CLIA & THSTI-RBD ELISA. Total true positive samples evaluated: 379; true negative samples: 184

| Tests | Sensitivity (95%CI) | Specificity |
| --- | --- | --- |
| Zydus-Kavach | 75.7 (71.0 - 79.9) | 99.5% |
| DiaSorin CLIA | 82.6 (78.3 - 86.2) | --- |
| RBD ELISA | 84.7 (80.6 - 88.1) | 100% |

**Figure 1.**
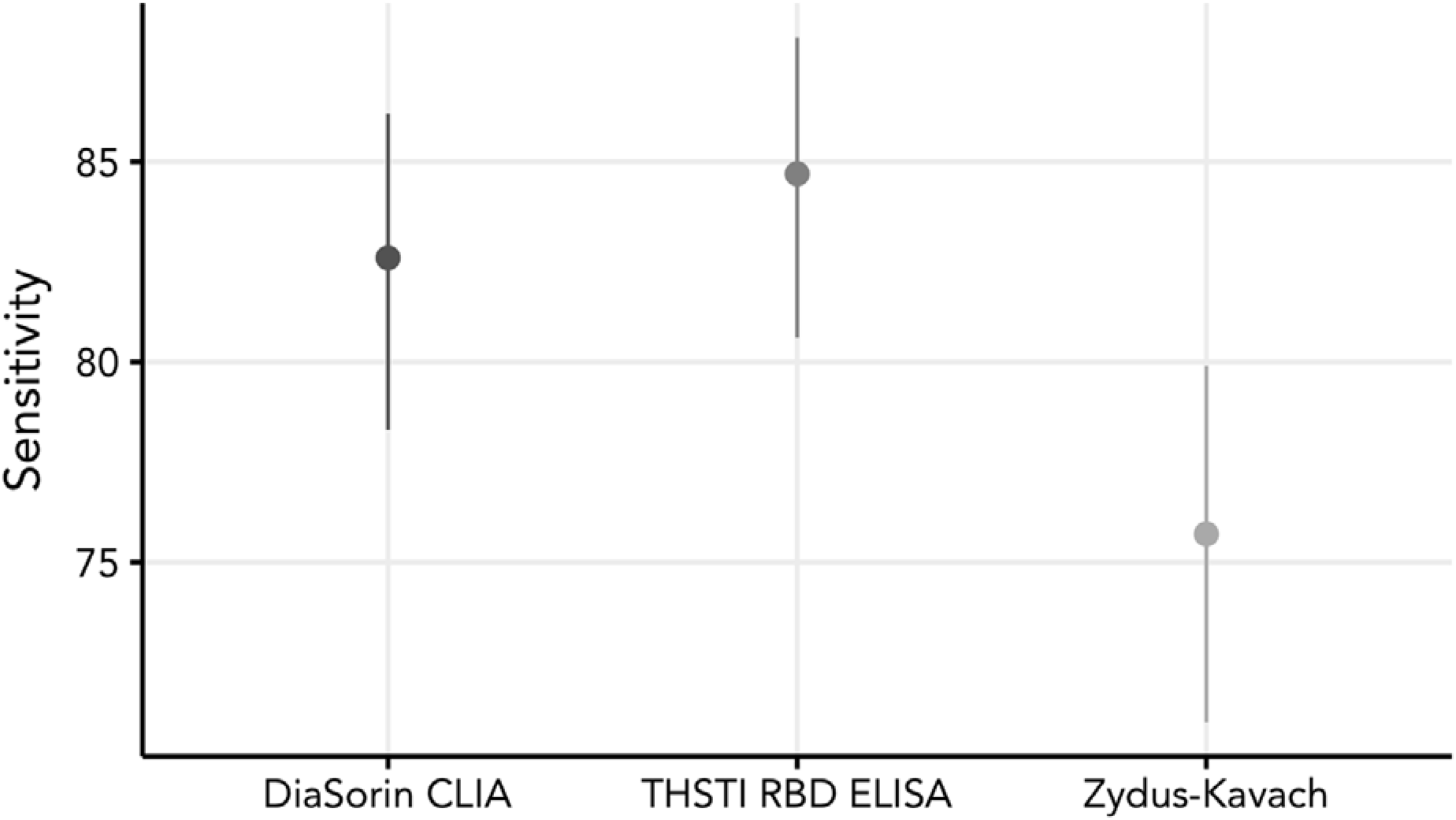
Plot of sensitivity (95% confidence intervals) for the candidate assays. The dots represent the sensitivity (%) and the bars, 95% CI.

The specificity of DiaSorin could not be evaluated due to limited availability of pre-pandemic negative sera.

### Comparison of Zydus-Kavach, DiaSorin CLIA & RBD ELISA

When evaluated for agreement between the tests, DiaSorin CLIA and RBD ELISA had highest concordance with a global agreement of 94.7% (95%CI: 92.0, 96.6). There was minimal bias between the two tests; with just 6 samples positive with DiaSorin labelled negative by RBD ELISA. Among the other 14 discordant samples that were positive by RBD ELISA, five were equivocal by DiaSorin and the rest were negative. The agreement estimated by prevalence and bias-adjusted kappa statistic (0.89) was near perfect between the two tests (Table-2 & Supplementary table-4).

On the other hand, head-to-head comparison of DiaSorin CLIA and RBD ELISA against Zydus Kavach demonstrated that the degrees of agreement were modest. The global agreement between the pairs of DiaSorin CLIA and Zydus Kavach, and RBD ELISA and Zydus Kavach were 88.7% (95%CI: 85.1, 91.5) and 87.3% (95%CI: 83.6, 90.3) respectively (Table-2). DiaSorin CLIA and RBD ELISA were superior to Zydus Kavach ELISA and were able to correctly identify 7% and 9% more IgG positive sera than the latter (Supplementary tables 2 & 3).

**Table 2:** Head-to-head comparison of Zydus-Kavach, DiaSorin CLIA & RBD ELISA.

| ***Kit*** | ***Zydus-Kavach*** | ***DiaSorin CLIA*** | ***RBD ELISA*** |
| --- | --- | --- | --- |
| ***Zydus-Kavach*** |  | ***88.7% (95%CI: 85.1, 91.5)*** | ***87.3% (95%CI: 83.6, 90.3)*** |
| ***DiaSorin CLIA*** | ***0.77*** |  | ***94.7% (95%CI: 92.0, 96.6)*** |
| ***RBD ELISA*** | ***0.75*** | ***0.89*** |  |

Numbers in the cells below the diagonal in the table denote Prevalence and Bias adjusted Kappa statistic. Numbers in the cells above the diagonal in the table denote agreement and 95%CI calculated as (positive by both methods + negative by both)/ total samples

## Discussion

Comparison of three assays on a well-characterized sample set showed that the DiaSorin CLIA and the in-house RBD ELISA performed slightly better than the Zydus Kavach ELISA with higher sensitivity and ability to identify more IgG positive samples. These results are important as they help interpretation of the serosurveillance studies that are being conducted in India using these tests.

The sensitivities reported in our study are less than reported by the companies or elsewhere [5,7]. An independent evaluation of DiaSorin CLIA, the sensitivity was reported to be 95.0% (95%CI: 92.8, 96.7), which is at least 13 percentage points higher than this report [3]. While unlikely, this difference could be attributed to false positives in our RT-PCR assay since we used RT-PCR positive convalescent samples as reference standards. Similarly, the article reporting internal validation of Zydus Kavach ELISA reported a sensitivity of 92.37% based on samples that were positive in a micro-neutralization assay as against 75.7% that we see in our study using sera collected >20 days post-RT-PCR positivity [7]. While we harmonised our definition of positive reference standard with that of the UK MHRA, about 14% of our participants were asymptomatic, and may have had a lower antibody response as reported in a longitudinal study, albeit with small numbers, which showed that 2/24 of their participants did not seroconvert when followed up 20-28 days into their illness [11].

To overcome the limitation of an imperfect positive reference standard and to improve inferences, the relative performance of these tests was evaluated by head-to-head comparison. RBD ELISA and DiaSorin CLIA were able to identify more positive IgG sera/plasma than Zydus Kavach. However, RBD ELISA is an in-house ELISA developed using similar sample collections. While the sample panel used for evaluation was independent of that used in the development of the RBD ELISA, its true test would be when it is evaluated externally.

Highly sensitive serological assays can assess immune response to SARS-CoV-2 and are needed to determine the extent of spread of the virus, which in turn is critical for assessing case fatality rates and herd immunity. Serological assays also help in assessing development of herd immunity to devise community management strategies that are of crucial importance at this time, and will continue to be relevant in the coming years. Other uses of serological assays can be to assess exposure in high-risk populations such as healthcare workers and assess vaccination strategy at state or national level. Cross-reactivity with SARS-CoV and seasonal coronaviruses in different population and timing of IgM and IgG responses need to continue to be considered. Till date, studies on comparative performance serological assays for SARS-CoV-2 show a range of sensitivity of 84-98% and specificity of 96-99% [12-29]. Two of the three IgG assays in this study used the Spike protein (Receptor binding domain or RBD in THSTI-In-house ELISA and S1/S2 in DiaSorin CLIA), while the specific antigenic epitope(s) of the inactivated virus in the Kavach assay are not defined. Since RBD in the spike protein is the major site of ACE-2 binding, assays with this target may have more concordance with neutralizing antibodies. The spike protein has more CD4 and CD8 T cell immunodominant epitopes as experimentally shown in SARS-CoV, and since these epitopes are mostly conserved even in SARS-CoV-2 isolates, serological assays targeting RBD or full length S1/S2 are putatively more appropriate for assessing long-term immune status [30].

## Conclusion

Overall, this is the first comparative study of SARS-CoV-2 IgG assays in India evaluated independently against a strategically designated alternate reference standard. One limitation of this study is that we were unable to evaluate the specificity of the Diasorin assay due to paucity of negative samples. Nonetheless, the well characterized panels provide useful information for decision-making for choice of serological assays.

## Data Availability

Data will be available on request to the corresponding author

**Abbreviations**
RBD: Receptor Binding Domain
CLIA: Chemi-Luminescence Immuno Assay
ELISA: Enzyme Linked ImmunoSorbent Assay
MHRA: The UK Medicines and Healthcare products Regulatory Agency

Writing committee:

Susmita Chaudhuri^*^, Ramachandran Thiruvengadam^*^, Pallavi Kshetrapal, Gaurav Batra, Tripti Shrivastava, Bapu Koundinya Desiraju, Gagandeep Kang^$^, Shinjini Bhatnagar#

* Equally contributed to the manuscript

$ Co-corresponding author

\# Corresponding authors

## Members of DBT India consortium for COVID-19 Research

Coordinating Institute: Translational Health Science and Technology Institute (THSTI)

Coordinating Principal Investigator: Dr Shinjini Bhatnagar

Co-Principal Investigator: Dr Gagandeep Kang

Co-Investigators (Clinical): Drs Nitya Wadhwa, Uma Chandramouli Natchu, Ramachandran Thiruvengadam, Shailaja Sopory, Pallavi Kshetrapal, Bapu Koundinya Desiraju, Vandita Bhartia, Mudita Gosain

Co-Investigators (Laboratory): Drs Gaurav Batra, Guruprasad Medigeshi, Susmita Chaudhuri, Niraj Kumar, Tarun Sharma, Chandresh Sharma, Shailendra Mani, Tripti Shrivastava

Clinical Operations Lead: Dr. Monika Bahl

International Centre for Genetic Engineering and Biotechnology (ICGEB): Dr Anmol Chandele

Delhi University, South Campus, New Delhi: Dr Vijay Kumar Chaudhary

National Institute of immunology (NII): Drs. Amulya Panda, Nimesh Gupta

Maulana Azad Medical College and Lok Nayak Jai Prakash Narayan Hospital, New Delhi: Drs. Nandini Sharma, Pragya Sharma, Sonal Saxena, J.C. Passey, Suresh Kumar

ESI Medical College and Hospital, Faridabad, Haryana: Dr Asim Das, Anil K Pandey, Nikhil Verma

Civil Hospital Gurugram (GCH), Haryana: Drs Deepa Sindhu, Jai Singh Malik

Civil Hospital Palwal (PCH), Haryana: Dr Brahmdeep Sindhu

Al-Falah School of Medical Science & Research Centre and Hospital, Dhouj, Haryana: Drs S.K.S. Puri, Bhupinder Kaur Anand, Shubham Girdhar

Medanta Hospital, Gurugram, Haryana: Drs Sushila Kataria, Pooja Sharma

Shaheed Hasan Khan Mewati Government Medical College, Nalhar, Nuh, Haryana: Dr Yamini

Lady Hardinge Medical College, New Delhi: Drs. Harish K. Pemde, Tanmaya Talukdar SGT Medical college, Gurugram, Haryana: Drs. Pankaj Abrol, Mukesh Sharma Dr. Dang’s Lab, New Delhi: Drs Navin Dang, Manavi Dang, Arjun Dang, Leena Chatterjee, Devjani De

## Author contribution

SB, GK, SC and RT conceptualized the study; All members of DBT India consortium for COVID-19 Research devised methods, collected samples and clinical data; SC, GB, TS, PK, ND, MD, AD, LC and DD provided reagents and analysed samples; RT, SC and BKD curated and analysed data; SC, RT and GK drafted the manuscript; GK and SB coordinated and supervised the study. All authors contributed in revision and approved the final draft of the manuscript.

## Acknowledgement

We deeply thank the Department of Biotechnology, Government of India for supporting the consortium. We are grateful to the leadership and administration of all partner institutions in the consortium for their help and support. We thank all the clinical, laboratory and data management staff for their contributions to this work and the consortium at large.

## Supplementary material

**sTable 1:**
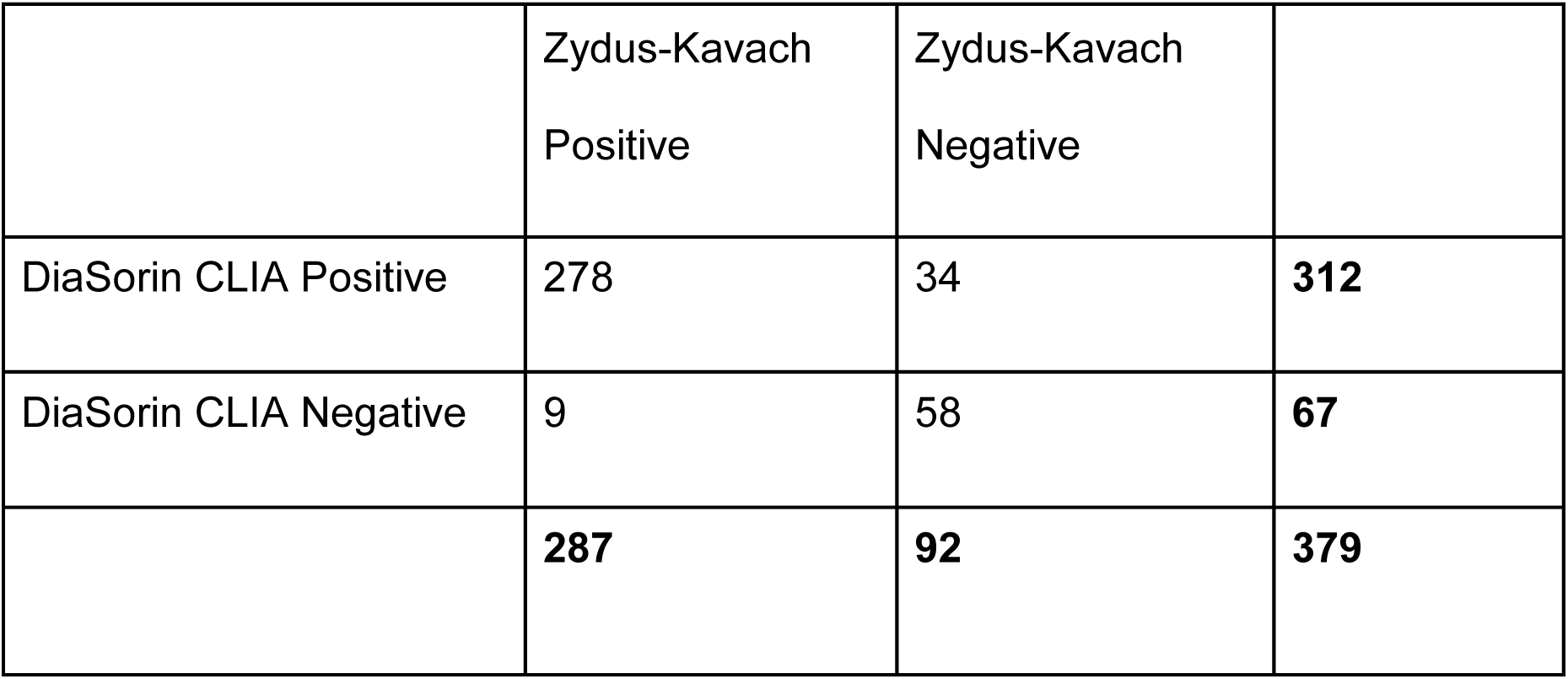
Head-head comparison of Zydus-Kavach & DiaSorin CLIA.

|  | Zydus-Kavach<br>Positive | Zydus-Kavach<br>Negative |  |
| --- | --- | --- | --- |
| DiaSorin CLIA Positive | 278 | 34 | **312** |
| DiaSorin CLIA Negative | 9 | 58 | **67** |
|  | **287** | **92** | **379** |

**Agreement/concordance analysis**

Global agreement: 88.7% (95%CI: 85.1, 91.5)

Specific agreement for positivity: 92.8% (95%CI: 90.5, 94.6)

Specific agreement for negativity: 73.0% (95%CI: 65.6, 79.3)

Cohen’s Kappa statistic: 0.66 (95%CI: 0.57, 0.75)

**Observed disagreement:**

DiaSorin+Kavach-: 8.97% Kavach+DiaSorin-: 2.37%

Bias index (Difference in the proportion of positivity)= 0.07

DiaSorin is able to identify more cases than Zydus Kavach

Prevalence index: 0.58

**Prevalence and Bias-adjusted Kappa (PABAK): 0.77** (Inference: *Substantial* agreement)

**sTable 2:** Head-head comparison of Zydus-Kavach & RBD ELISA.

|  | Zydus-Kavach<br>Positive | Zydus-Kavach<br>Negative |  |
| --- | --- | --- | --- |
| RBD ELISA positive | 280 | 41 | **321** |
| RBD ELISA Negative | 7 | 51 | **58** |
|  | **287** | **92** | **379** |

**Agreement/concordance analysis**

Global agreement: 87.3% (95%CI: 83.6, 90.3)

Specific agreement for positivity: 92.1% (95%CI: 89.7, 94.0)

Specific agreement for negativity: 68.0% (95%CI: 60.2, 74.9)

Cohen’s Kappa statistic: 0.61 (95%CI: 0.51, 0.70)

**Observed disagreement:**

RBD ELISA+Kavach-: 10.8% Kavach+RBD ELISA: 1.9%

Bias index (Difference in the proportion of positivity)= 0.09

RBD ELISA is able to identify more cases than Zydus Kavach

Prevalence index: 0.60

**Prevalence and Bias-adjusted Kappa (PABAK): 0.75** (Inference: *Substantial* agreement)

**sTable 3:** Head-head comparison of DiaSorin & RBD ELISA.

|  | **DiaSorin**<br>**Positive** | **DiaSorin**<br>**Negative** |  |
| --- | --- | --- | --- |
| **RBD ELISA positive** | 307 | 14 | **321** |
| **RBD ELISA negative** | 6 | 52 | **58** |
|  | **313** | **66** | **379** |

**Agreement/concordance analysis**

Global agreement: 94.7% (95%CI: 92.0, 96.6)

Specific agreement for positivity: 96.8% (95%CI: 95.2, 97.9)

Specific agreement for negativity: 83.9% (95%CI: 76.4, 89.3)

Cohen’s Kappa statistic: 0.81 (95%CI: 0.73, 0.89)

**Observed disagreement:**

RBD ELISA+DiaSorin-: 3.7% DiaSorin+RBD ELISA: 1.6%

Bias index (Difference in the proportion of positivity)= 0.02

Minimal bias between the RBD ELISA and DiaSorin

Prevalence index: 0.67

**Prevalence and Bias-adjusted Kappa (PABAK): 0.89** (Inference: *Almost perfect* agreement)

**sTable 4:** Sensitivity of Zydus Kavach, DiaSorin, RBD ELISA across various periods of illness.

| Duration of illness (days) | N | Zydus Kavach |  | DiaSorin CLIA |  | RBD ELISA |  |
| --- | --- | --- | --- | --- | --- | --- | --- |
|  |  | n | Sensitivity (%) | n | Sensitivity (%) | n | Sensitivity (%) |
| 20-30 | 193 | 145 | 75.1 | 163 | 84.5 | 167 | 86.5 |
| 31-40 | 6 | 6 | 100 | 6 | 100 | 5 | 83.3 |
| 41-50 | 116 | 83 | 71.6 | 89 | 76.7 | 92 | 79.3 |
| 51-60 | 56 | 48 | 85.7 | 50 | 89.3 | 51 | 91.1 |
| 61-72 | 4 | 2 | 50 | 2 | 50 | 3 | 75.0 |

**sFigure 1:**
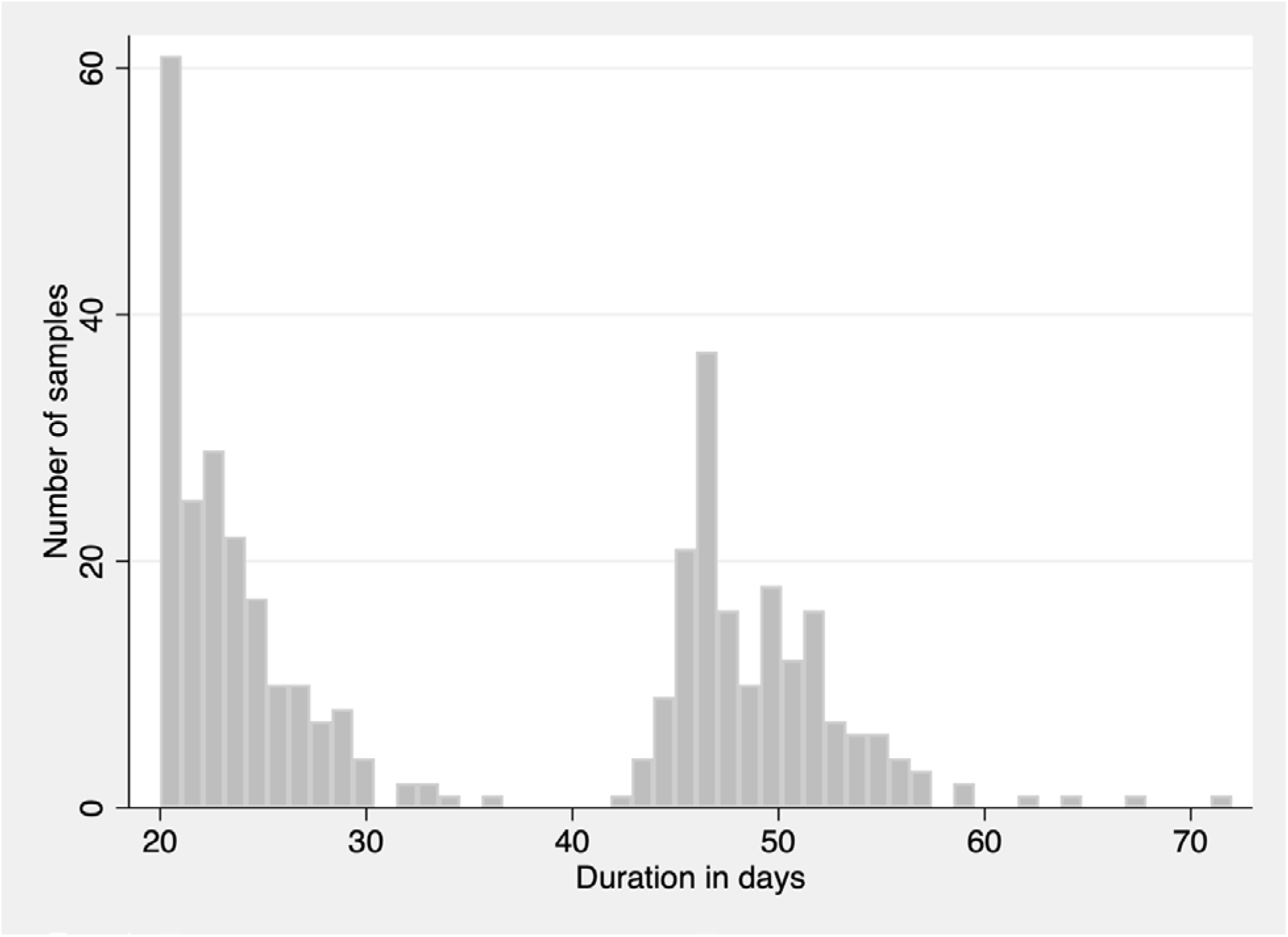
Number of samples over a range of duration of illness.

